# PRERISK: A Personalized, daily and AI-based stroke recurrence predictor for patient awareness and treatment compliance

**DOI:** 10.1101/2023.03.24.23287721

**Authors:** Giorgio Colangelo, Marc Ribo, Estefania Montiel, Didier Dominguez, Marta Olive, Marian Muchada, Alvaro Garcia-Tornel, Manuel Requena, Jorge Pagola, Jesus Juega, David Rodriguez-Luna, Noelia Rodriguez-Villatoro, Federica Rizzo, Belen Taborda, Carlos A. Molina, Marta Rubiera

**Affiliations:** VHIR - Vall d’Hebron Institute of Research, Passeig de la Vall d’Hebron, 129, 08035 Barcelona; Hospital Universitari Vall d’Hebron, Stroke Unit, Neurology Department, Passeig de la Vall d’Hebron, 129, 08035 Barcelona; Nora Health, Passeig de la Vall d’Hebron, 129, 08035 Barcelona; Programa d’Analítica de Dades per a la Recerca i la Innovació en Salut (PADRIS), Agència de Qualitat i Avaluació Sanitàries de Catalunya (AQuAS), Departament de Salut | Generalitat de Catalunya, Carrer de Roc Boronat, 81-95 | 08005 Barcelona

## Abstract

**BACKGROUND:** The risk prediction of stroke recurrence for individual patients is a difficult task. Individualised prediction may enhance stroke survivors selfcare engagement. We have developed PRERISK: a statistical and Machine Learning (ML) classifier to predict individual stroke recurrence risk.

**METHODS:** We analysed clinical and socioeconomic data from a prospectively collected public healthcare-based dataset of 44623 patients admitted with stroke diagnosis in 88 public hospitals over 6 years in Catalonia-Spain. We trained several supervised-ML models to provide individualised risk along time and compared them with a Cox regression model.

**RESULTS:** Overall, 16% of patients presented a stroke recurrence along a median follow-up of 2.65 years. Models were trained for predicting early, late and long-term recurrence risk, within 90, 91-365 and >365 days, respectively. Most powerful predictors of stroke recurrence were time since index stroke, Barthel index, atrial fibrillation, dyslipidemia, haemoglobin and body mass index, which were used to create a simplified model with similar performance. The balanced AUROC were 0.77 (±0.01), 0.61 (±0.01) and 0.71 (±0.01) for early, late and long-term recurrence risk respectively (Cox risk class probability: 0.74(±0.01), 0.59(±0.01) and 0.68(±0.01), c-index 0.88). Overall, the ML approach showed statistically significant improvement over the Cox model. Stroke recurrence curves can be simulated for each patient under different degrees of control of modifiable factors.

**CONCLUSION:** PRERISK represents a novel approach that provides continuous, personalised and fairly accurate risk prediction of stroke recurrence along time according to the degree of modifiable risk factors control.

**CLINICAL PERSPECTIVE:** *What is new?:*

- Stroke recurrence is frequent after stroke despite advances in stroke treatments, and it is difficult to predict the individual risk of one patient.
- We have created PRERISK, a predictive model based on machine learning (ML) which provides individualised information of the probability of stroke recurrence and can be re-calculated according to risk factors control.

*What are the clinical implications?:*

- PRERISK information can be used as feedback for secondary prevention strategies and enhance patient engagement and treatment compliance.
- It could be scalable to optimise ML-based prevention strategies in other chronic conditions.

## INTRODUCTION

Worldwide 1 out of 4 strokes are recurrences, implying about three million events each year ^1,2^. The rate of recurrence after a first ever stroke ranges from 16 to 30% at 5 years in different series ^3–5^. A second stroke has been associated with longer in-hospital stay, increased mortality, and a higher degree of disability ^6,7^. In young patients, a recurrent stroke increases mortality by 14 times as compared with the first ischemic event, and by 30 times compared with the age-adjusted general population death rate ^8^. Compliance with guideline-based secondary stroke prevention has demonstrated to reduce stroke recurrence (8). Therefore, over the last years substantial attempts have been made to increase guidelines compliance and prevent recurrences. Despite all these efforts, recent epidemiological studies have not found a sustained trend in the reduction of stroke recurrence rate, highlighting a non-resolved healthcare problem ^3,4^.

Ischemic stroke and its recurrence are multifactorial conditions influenced by a combination of genetic, environmental, and vascular factors. Some of them are related to the stroke aetiology or mechanism (as atherothrombotic aetiology, atrial fibrillation, etc) ^9,10^. Several risk factors such as age are non-modifiable; however, other factors like treatment compliance or sedentary lifestyle can be modified by the patient’s active participation in the secondary prevention strategies. A recent study showed that more than 90% of the stroke burden is attributable to modifiable risk factors and achieving a control of behavioural and metabolic risk factors could avert more than 75% of the global stroke burden ^11^. However, despite the known role of risk factors in stroke recurrence, it is difficult to individualise the specific risk of stroke recurrence for each patient along time.

Several scores based on proportional hazard models have been developed aiming to predict the individual stroke recurrence risk in specific populations ^12–14^. However, these scores often require adding advanced imaging information such as MRI data to increase their accuracy, which restricts its generalizability worldwide ^15^ or are tested on small cohorts.

In the last few years, machine learning (ML) has emerged as a valuable tool to address several health problems ^16,17^ including cardiovascular diseases ^18^ and stroke ^19,20^. It allows the analysis of several factors, depicting interactions that may not be easy to predict based on previous knowledge or to identify with classical statistical analysis.

In the present study, we aimed to generate PRERISK: a transparent, easy to acquire, individual predictive model for stroke recurrence risk at different time-points based on ML and statistical models. We also compared the ML model with a classical time varying proportional hazard model. The individual prediction for each patient is aimed to be dynamic, allowing re-calculation over time according to the degree of control of the modifiable risk factors included in the model. Therefore, a continuous estimation of the risk reduction if these factors are well controlled is provided.

## PATIENTS AND METHODS

A clinical and socioeconomic public healthcare-based dataset of 44623 patients admitted with stroke diagnosis in 88 public hospitals in Catalonia over 6 years (2014-2020) was analysed. Anonymized data were provided by the Catalan Agency for Health Quality and Evaluation (AQUAS) within the framework of the Data Analytics Program for Health Research and Innovation (PADRIS). The PADRIS allows access to information from different sources on public healthcare resources usage for the population of Catalonia linked at the patient level with warranted accomplishment of ethical principles. Patients were extracted based on the International Statistical Classification of Diseases and Related Health Problems (ICD) 9 and 10 stroke-related codes (see Online Supplementary Material, Table I), and were classified as ischemic stroke (IS), intracranial haemorrhage (ICH) or transient ischemic attack (TIA). 36521 patients were selected with a first IS or ICH, as represented in Figure 1A. Patients who died within 7 days from the index stroke (first event or recurrence) were excluded from the analysis. Stroke recurrences recorded within 1 day from the index event were considered as clinical fluctuations of the index stroke and therefore not considered as a new event.

**Figure 1:**
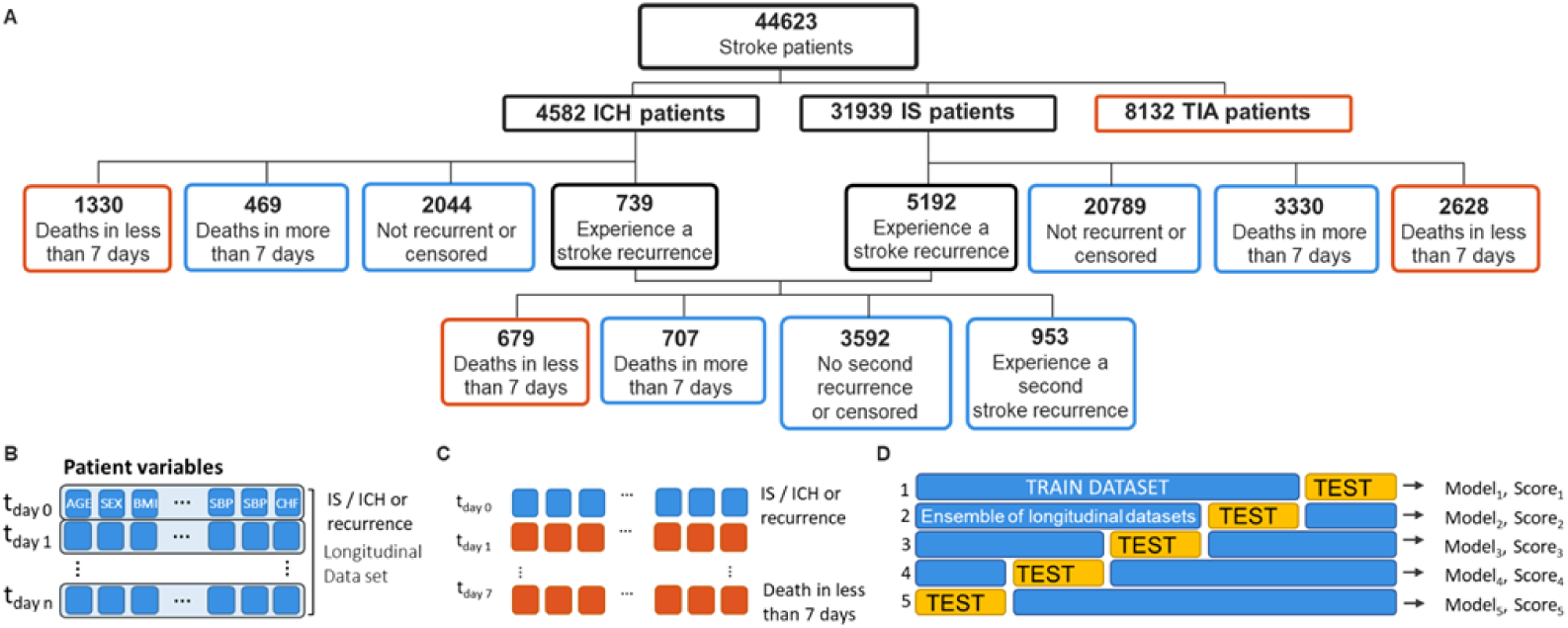
**A)** Included patient cohort diagram. Blue and black boxes represent patients included in the study, red boxes discarded patients. Longitudinal data from the last stroke to death were excluded from the analysis if death occurred within 7 days. Stroke recurrences recorded within 1 day from the index event were considered as clinical fluctuations of the index event and therefore not considered as a new event. ICH: intracranial haemorrhage; IS: ischemic stroke. **B-C)** Inclusion scheme for longitudinal data sets for a single patient. Red and Blue boxes represent single variables at multiple times t: t_day 0_, , … t_day N_. Blue / red boxes represent data used / excluded for the model training. Index event: IS, ICH or recurrence. BMI: body mass index; SBP: systolic blood pressure; CHF: cardiac heart failure **D)** 5-fold cross validation scheme used in this work. The train and test datasets are made with sets at t_n_ of all the patients. The global data set of data points included in the model are splitted in two independent train and test data sets (with approximate ratio of 4 to 1), with different patients between train and test data sets. Each train data set is used to train a model and the test data set is used to validate its accuracy. The same procedure is repeated using different patients across the overall data set. Finally models calculated in each training session are compared and an overall performance is calculated.

The complete set of variables (Table 1) was created integrating variables belonging to different datasets: admissions, pharmacy data, laboratory analysis, home-related socio-economic data, etc. To remark, no personal social or economic data were included in the analysis: the district socio-economic level is an index ranging from 0 to 100 that refers to the available social facilities in the patients’ district (primary health centres, public schools, etc) ^21^. With the information provided in the collected datasets, we constructed other variables which provide practical patient indicators which can be easily exploited in the models: the “Emergency admissions in the last 3 months” is a binary variable describing whether the patient had an urgent admission within the three months before the stroke event, and it discriminates between admission caused by a stroke or any other pathology.

**Table 1:** Baseline clinical characteristics of the series, ordered by P-values. P-values are calculated on the distributions with patients with and without stroke recurrence within 365 days. Patients with intrahospital death (within 7 days from the stroke event) are excluded from the baseline calculation). *: variables derived from previous codified diagnosis according to the International Statistical Classification of Diseases and Related Health Problems (ICD) 9 and 10; ICD codes associated with variables are reported in Table I of supplementary information. Patients without an ICD code were assumed to be not affected. IQR: interquartile range.

|  | Not<br>Recurrent | Recurrent | IQR<br>Not<br>Rec | IQR<br>Rec | P<br>value | %<br>Missing<br>Values |
| --- | --- | --- | --- | --- | --- | --- |
| **Age (years)\*** | 77.5 | 77.5 | 20 | 20 | <0.00<br>1 | 0 |
| **Basic daily activities (Barthel)** | 69.76 | 65 | 51.77 | 51.1<br>2 | <0.00<br>1 | 50.78 |
| **Body Mass Index (BMI)**<br>**(Kg/m2)** | 27.64 | 27.54 | 5.95 | 6.07 | <0.00<br>1 | 8.8 |
| **Creatinine (mg/dl)** | 0.92 | 0.92 | 0.36 | 0.37 | <0.00<br>1 | 4.75 |
| **Diastolic blood pressure**<br>**(DBP) (mmHg)** | 74.24 | 74.17 | 11.31 | 12.2<br>8 | <0.00<br>1 | 5.36 |
| **District socio-economic level**<br>**(0-100)** | 58.98 | 58.01 | 17.26 | 15.4<br>5 | <0.00<br>1 | 1.49 |
| **Glycaemia (mg/dl)** | 102.81 | 104.74 | 35.97 | 40 | <0.00<br>1 | 4.7 |
| **Haemoglobin A1c (HbA1c) (%)** | 5.63 | 5.79 | 1.64 | 1.77 | <0.00<br>1 | 30.86 |
| **Height (cm)** | 162 | 162 | 15 | 14 | <0.00<br>1 | 12.06 |
| **Haemoglobin (g/dl)** | 13.46 | 13.39 | 2.37 | 2.32 | <0.00<br>1 | 4.94 |
| **Low-income district** | 1 | 1.01 | 0.18 | 0.16 | <0.00<br>1 | 1.49 |
| **Past strokes (Number)\*** | 1 | 2 | 0 | 1 | <0.00<br>1 | 0 |
| **Systolic blood pressure (SBP) (mmHg)** | 131.23 | 131.64 | 15.25 | 17.1<br>6 | <0.00<br>1 | 5.36 |
| **Time from previous stroke (days)** | 933 | 144 | 975.2<br>5 | 814 | <0.00<br>1 | 0 |
| **Total cholesterol (mg/dl)** | 163.41 | 163.87 | 50.68 | 52.4<br>3 | <0.00<br>1 | 5.38 |
| **Alcohol abuse (Y)\*** | 12.29<br>% | 16.95<br>% | - | - | <0.00<br>1 | 0 |
| **Anaemia (Y)\*** | 20.59<br>% | 24.4 % | - | - | <0.00<br>1 | 0 |
| **Anticoagulant treatment (Y)** | 25.89<br>% | 29.6 % | - | - | <0.00<br>1 | 3.76 |
| **Antidepressants (Y)** | 24.98<br>% | 24.41<br>% | - | - | <0.00<br>1 | 0.63 |
| **Antidiabetics (Y)** | 23.58<br>% | 25.32<br>% | - | - | <0.00<br>1 | 0.63 |
| **Antiepileptic drugs (Y)** | 12.77<br>% | 15.17<br>% | - | - | <0.00<br>1 | 0.63 |
| **Antihypertensives (Y)** | 62.03<br>% | 62.85<br>% | - | - | 0.011<br>2 | 0.63 |
| **Antiplatelet treatment (Y)** | 70.53<br>% | 66.06<br>% | - | - | <0.00<br>1 | 3.76 |
| **Anxiolytics (Y)** | 24.61<br>% | 24.99<br>% | - | - | <0.00<br>1 | 0.63 |
| **Anxious-depressive disorder (Y)\*** | 19.13 % | 22.45 % | - | - | <0.001 | 0 |
| **Atrial fibrillation (Y)\*** | 30.1 % | 39.66 % | - | - | <0.001 | 0 |
| **Chronic heart failure (CHF) (Y)\*** | 17.08 % | 21.01 % | - | - | <0.001 | 0 |
| **Chronic kidney disease (CKD) (Y)\*** | 19.06 % | 22.11 % | - | - | <0.001 | 0 |
| **Chronic obstructive pulmonary disease (COPD) (Y)\*** | 11.85 % | 14.74 % | - | - | <0.001 | 0 |
| **Coronary heart disease (Y)\*** | 14.85 % | 18.8 % | - | - | <0.001 | 0 |
| **DBP>80 (Y)\*** | 22.4 % | 22.6 % | - | - | <0.001 | 0 |
| **Diabetes Mellitus (Y)\*** | 37.06 % | 40.93 % | - | - | <0.001 | 0 |
| **Dyslipidemia (Y)\*** | 59.02 % | 66.84 % | - | - | <0.001 | 0 |
| **Emergency in last 3M (stroke)** | 7.77 % | 46.61 % | - | - | <0.001 | 0 |
| **Emergency in last 3M (no stroke)** | 13.04 % | 16.01 % | - | - | <0.001 | 0 |
| **Sex (Female)** | 45.39 % | 44.83 % | - | - | <0.001 | 0 |
| **HbA1c >7 (Y)\*** | 10.46 % | 11.89 % | - | - | <0.001 | 0 |
| **High blood Pressure (HBP) (Y)\*** | 77.43 % | 83.04 % | - | - | <0.001 | 0 |
| **Obesity (Y)\*** | 14.83 % | 16.77 % | - | - | <0.001 | 0 |
| **Past intracranial haemorrhage (ICH) (Y)\*** | 9.44 % | 14.38 % | - | - | <0.001 | 0 |
| **Past ischemic stroke (IS) (Y)\*** | 90.56 % | 89.02 % | - | - | <0.001 | 0 |
| **Past transient ischemic attack (TIA) (Y)\*** | 5.01 % | 4.86 % | - | - | <0.001 | 0 |
| **SBP>130 (Y)** | 45.03 % | 44.98 % | - | - | <0.001 | 0 |
| **Statins (Y)\*** | 51.51 % | 48.99 % | - | - | 0.0075 | 0.63 |
| **Tobacco addiction (Y)** | 38.98 % | 41.81 % | - | - | <0.001 | 0 |

For each patient, time was set to zero at the index event (first stroke or recurrence). From the index event, patient variables are associated to subsequent time steps (days) where information was collected. Variables were linearly interpolated to fill missing values if information for that time step was not available. When not interpolable, missing values of the included variables were completed with the mode of the dataset. Missing values of “Basic daily activities (Barthel scale)” were completed with the mean value of patients with the same age, and with 100 for patients younger than 75 years. The same procedure was repeated independently for both the train and the test datasets.

We trained the ML algorithms considering the “time to stroke recurrence” as target. We translated the problem of estimating the stroke recurrence risk into a classification problem: an “early risk” label was associated to sets describing the patient who is going to experience a stroke recurrence within 90 days, “late risk” to sets describing patients who are going to experience a stroke recurrence between 90 and 365 days, and “long term risk” to sets describing the patients who are not going to experience a stroke within the following 365 days. Censored patients with a tracking history of less than 365 days were not included in the ML training. In total 13320 entries were labelled as early risk, 10219 as late risk and 341396 as long-term risk. Figure 1B-D summarises the data selection schema for longitudinal data of each patient.

Different classification ML based models are analysed in this study: Random Forest (RF), AdaBoost (ADA) and Extreme Gradient Boosting (XGB). Our choice was motivated by the possibility of using most performant, widely adopted and documented models which can be easily reproduced by other clinical centres. Moreover, in these models explicability can be inferred by several computational methods (out of bag (OOB) in addition to permutation importance, which is used in this study to ease comparison with other studies). Grid search optimization has been performed for hyperparameter tuning. The classifiers are tested under different metrics: accuracy, F-score, recall, area under the receiver operator characteristics (AUROC); detailed information about other models and details about hyperparameters can be found in Table II-III of the Supplemental material. Because the original data set showed a strong imbalance among the number of early, late and long-term risk of stroke recurrence classes, we adjusted the weights of each class in the model training to be inversely proportional to the frequency of the class. To set a statistical benchmark for this analysis, we estimated the patient recurrence time using the Cox model ^22^, a proportional hazard model, a golden standard for survival analysis. The goodness to fit for the Cox model was evaluated by the concordance C-index. We tested our models with a 5-fold cross validation scheme, as shown in Figure 1D: data of the global dataset were splitted in 5 segments: 4 segments were used for the training and the other for the test. Independent patients were used for training and test data sets. 5 independent models were trained and tested in 5 separate runs and mean performance score and standard deviation are calculated. 5-fold optimization preserved class distribution within 3.5%, 1.3% and 0.1% tolerance for early, late and long recurrence classes respectively.

The present study has been approved by the local Ethics Committee (PR(AG)107/2020). Given the anonymous registry data, no informed consent was required. The data that support the findings of this study are available from the corresponding author, [MR], upon reasonable request.

## RESULTS

### Data Exploration

In total, for each patient, more than 40 variables were evaluated and summarised in Table 1. Median age of the series was 77.5 years, with 45% of females, and a high rate of vascular risk factors.

Figure 2 represents the distribution of the recurrences along the follow up time. From the 36521 patients with stroke diagnosis, 3958 died within 7 days from the index event, ending up in a cohort of 32563 patients.

**Figure 2:**
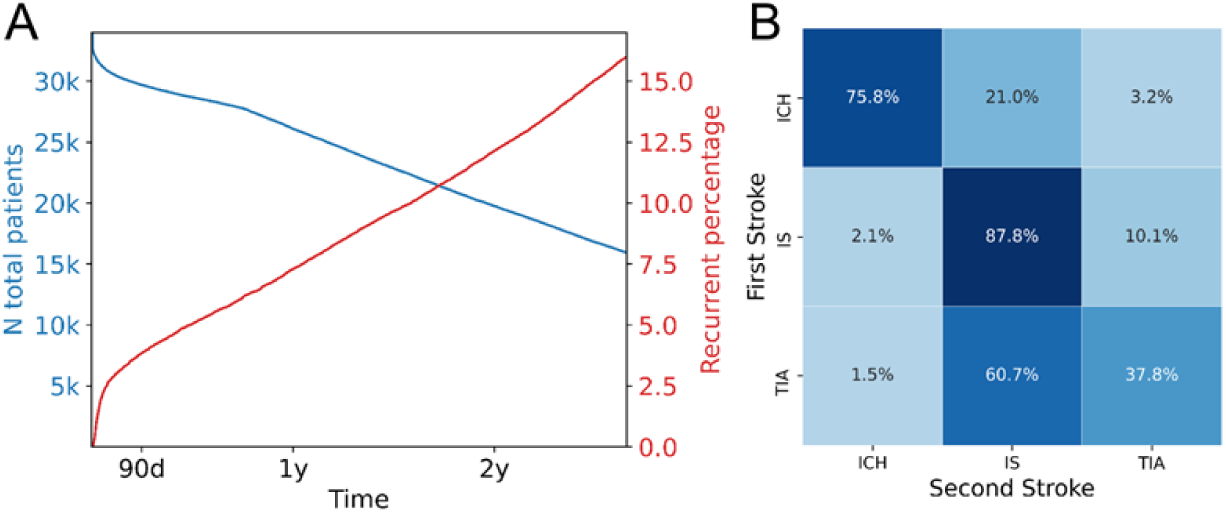
**A)** Red: Distribution of the recurrences of patients with ICH or IS along follow-up; blue: support curve. At day 0 the initial cohort was 36521 patients **B)** Stroke type recurrence matrix. In rows represented the type of first stroke (including TIA), in columns the percentage of the second event according to its type (IS, ICH, TIA). N: number. K: *1000; d: day; y: year; IS: ischemic stroke; ICH: intracranial haemorrhage; TIA: transient ischemic attack.

Of the restricted cohort, 5931 patients (16%) experienced a stroke recurrence (IS or ICH) along a median follow-up time of 2.65 years (IQR 2.74), and 953 patients (2.6%) experienced two or more stroke recurrences. Recurrences were generally of the same stroke subtype as the index stroke; however, up to 21% of patients with an index ICH experienced an ischemic recurrent stroke (Figure 2B). The recurrence rate was 3.85% at 90 days and 7.30% at 12 months.

### Models

We trained ADA, XGB and RF classifiers to predict the risk class for each patient at different time points. Figure 3 shows model performance in prediction of different recurrence classes. To overcome the high-class imbalance, we weighted each class by the support for each label. Both ADA and RF classifiers show improved area under the receiver operating characteristic (AUROC) with respect to statistical Cox models for all recurrence classes. No statistically significant improvement was observed for the XGB classifier.

**Figure 3:**
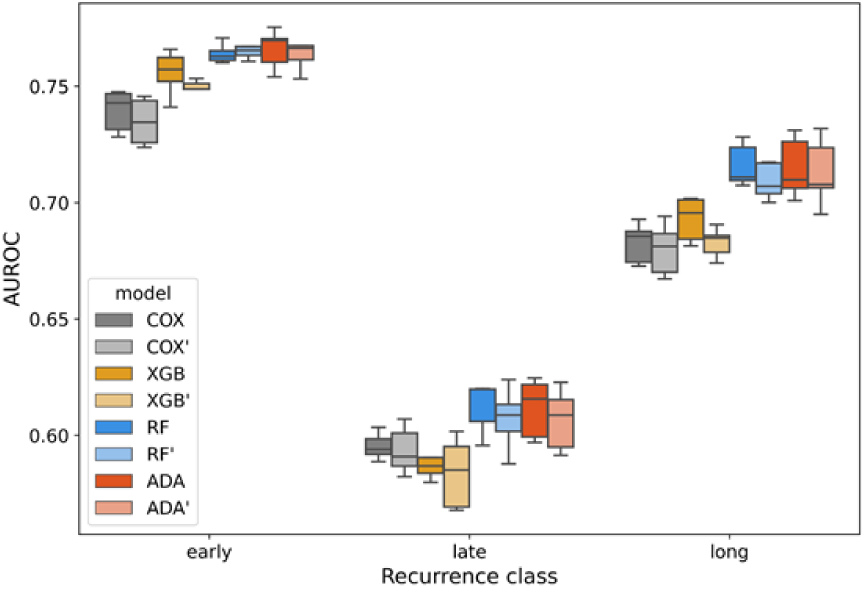
Area under the receiver operating characteristic (AUROC) comparison for early, late and long-term recurrence risk class for statistical (COX) and ML models (XGB, RF and ADA classifiers) with the full set of parameters (dark colours), or reduced set of parameters (identified with “‘”, light colours). A 5-fold cross validation test was used for the evaluation. RF: random forest; ADA: AdaBoost, XGB: Extreme Gradient Boosting.

The feature importance of the variables used in the model showed the most relevant predictive capability for atrial fibrillation, dyslipidemia, time from first stroke, haemoglobin, glycemia and age. Overall, modifiable cardiovascular risk factors account from 14% to 36% of permutation importance in different models (ADA: 14% (±2%), RF: 22% (±1%), 36(% (±1%), where number between parentheses represents standard deviations). The complete list of permutation importance for different models is represented in Figure 4A). We created an additional simplified model (ML’) using only the 7 most predictive features for ML model (Time from previous stroke, Barthel scale, Atrial fibrillation, Dyslipidemia, Age, Diabetes and Gender plus most of modifiable cardiovascular risk factors (Glycemia, Body Mass Index, High Blood Pressure, Cholesterol, Tobacco addiction, Alcohol abuse). The permutation importance of the included features is represented in Figure 4B. In the simplified ML’ model, modifiable cardiovascular risk factors accounted for 15% to 39% of the overall permutation feature importance of the model (ADA: 14% (±2%), RF: 23% (±1%), 38% (±1%)). As shown in Figure 4A-B, most performant ML models (ADA and RF), show similar qualitative permutation importance of the same features. Overall, although reduced ML’ models show lower accuracy than ML models, statistically significant improvement over COX model is observed for all the recurrence classes.

**Figure 4.**
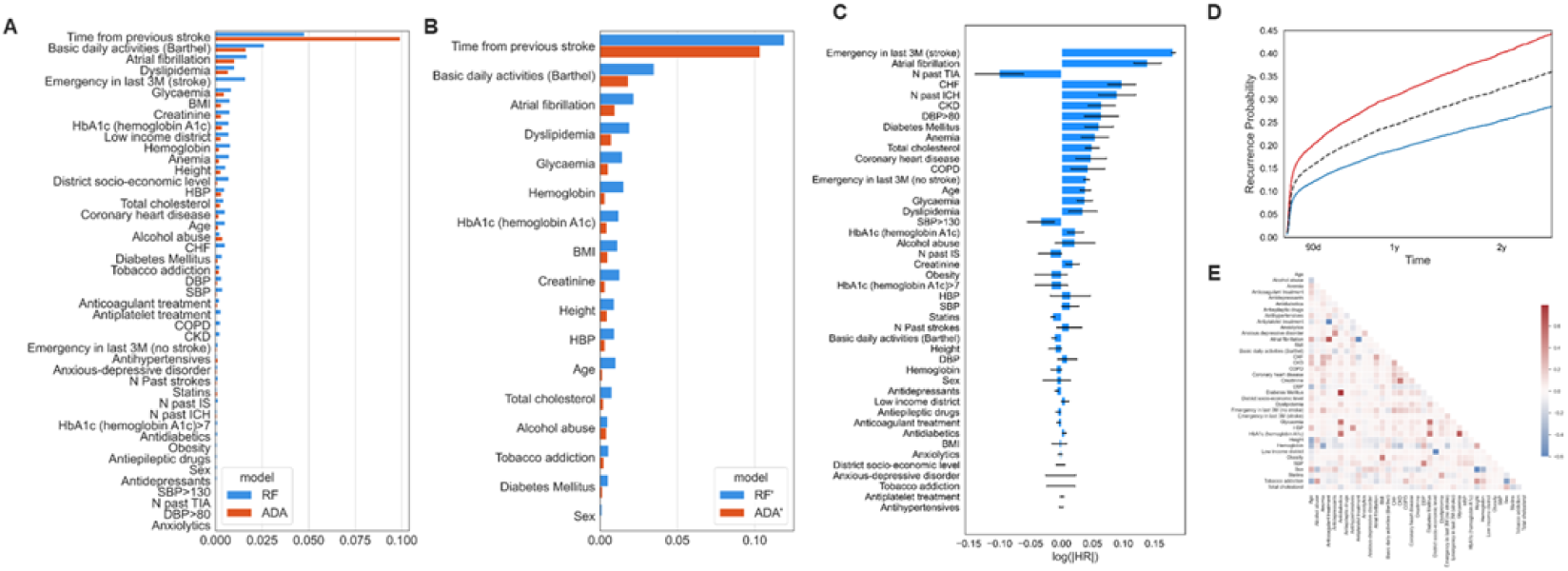
Permutation features importance of the two most performant models: RF, ADA classifiers when all available observables are used (A) and a reduced number of features is used to train the models RF’, ADA’ (B). RF: random forest; ADA: AdaBoost; BMI: body mass index; HBP: high blood pressure; DBP: diastolic blood pressure; CHF: chronic heart failure; SBP: systolic blood pressure; CKD: chronic kidney disease; COPD: chronic obstructive pulmonary disease; IS: ischemic stroke; ICH: intracranial haemorrhage; TIA: transient ischemic attack. **C)** Hazard ratios (HR) provided by Cox model for stroke recurrence. **D)** Example of stroke recurrence probability curves for the same patient with good (blue) and poor (red) control of cardiovascular risk factors: (Alcohol abuse, Body Mass Index, Creatinine, Cholesterol, Tobacco addiction, Glycaemia, blood pressure). Dark dashed line is average control of risk factors. Values used for “good” and “poor” control are based on two standard deviations below and above mean values. Ideal control of cardiovascular risk factors enable reduction of risk of recurrence by 28 % within 1 year with respect to mean values and 62% with respect to poor control scenarios. **E)** Correlation matrix of most relevant patient variables. BMI: body mass index; HBP: high blood pressure; DBP: diastolic blood pressure; CHF: chronic heart failure; SBP: systolic blood pressure; CKD: chronic kidney disease; COPD: chronic obstructive pulmonary disease; IS: ischemic stroke; ICH: intracranial haemorrhage; TIA: transient ischemic attack

We also analysed the hazard ratio of each variable using the statistical based Cox survival analysis. This approach enables evidence-based feedback to patients concerning his/her individual risk of recurrence. The dataset used for the Cox model was created aggregating longitudinal data lines for each patient, by average values, using information from one stroke to the following one. In total 10614 aggregated observations were used to train the model with 4021 events observed. The Cox model showed a c-index of 0.885. Complete Cox model estimates are reported in Table IV of Supplemental Material. Hazard Ratio Cox model offers a robust benchmark for personalised risk prediction for different values of one or more variables. The individual risk of a patient can be re-calculated according to risk factors control. Hazard Ratios provided by Cox model are represented in Figure 4C.

Figure 4D shows an example of an individual’s different survival curves according to good and poor modifiable risk factors control. However, this approximation does not take into account correlation among different variables. Correlations between the set of variables used for the simplified ML’ model are represented in Figure 4E.

Similarly, to ADA and RF classifiers, most powerful predictors in the Cox model were: a previous consultation in an emergency department in the 3 months prior to the index event and atrial fibrillation. However, Hazard Ratio scale and permutation importance are not directly comparable and for our purposes serve for different scopes.

We compared the performance of the ML model with the Cox regression model. As the Cox model provides a hazard probability *h(t)* that the patient experiences a recurrence, to directly compare the results with ML classifiers, we estimated the probability that a patient has a recurrence at time *t’* assigning early, late and long-term risk of recurrence as: *h(t’* ≤ *90)= h(90)*, *h(90*< *t’* ≤ *365)= (1-h(90))* (1-h(365)), h(t’* > *365)=1-h(365).* To compare the two approaches, we firstly trained the Cox model with aggregated data obtained by the mean values of each patient from one event to the other, then we calculated the risk class using a complete set of variables corresponding to the same time. The same set has been used to test the accuracy of the ML model. Complete and reduced set of variables was used to create COX and COX’ models respectively. Figure 3 shows the area under the curve (AUROC) for the three ML models (ADA, XGB and RF), in which independent patient data have been used for train and test sets. ML approaches (RF and ADA) shows statistical improvement over the Cox model for early (p<0.005) and long-term recurrence risk prediction (p<0.005) (mean AUROC=0.77 ±0.01, 0.61 ±0.01, 0.71 ±0.01, for ML model and AUROC = 0.74 ±0.01, 0.59 ±0.01, 0.68 ±0.01 for Cox model, for early, late and long-term risk of recurrence respectively), while less statistical difference was found in the two models for the late class recurrence risk (p-value = 0.033).

## DISCUSSION

We have developed PRERISK, a stroke recurrence predictive model based both on statistical and ML methods, which provides a fairly accurate prediction of stroke recurrence within the following 3 and 12 months after the first stroke. A simplified version of the models, focused on modifiable risk factors was also tested, providing almost equivalent results as compared to more complex models. These prediction tools can provide positive feedback to patients if modifiable vascular risk factors are adequately controlled.

Our model considered patients with all stroke subtypes: ischemic and hemorrhagic and could also be extended to TIA. As shown in Figure 2B, 98% of recurrent patients who experienced an IS as first event, experienced another ischemic event (IS or TIA) while only 2% had an ICH as recurrent event. Rates are different for recurrences in patients with an ICH as index event: 74% of them experienced another ICH and 21% an IS. Given the higher incidence of IS in the population, it is not uncommon to observe an IS recurrence in a patient with a previous ICH. Both ICH and IS share most vascular risk factors (age, high blood pressure, anticoagulation in atrial fibrillation, etc.); therefore, it is reasonable that a common predictive model performs well for both stroke subtypes.

PRERISK model provides accurate prognosis for early and long-term recurrences. Previous scores are usually restricted to short- or long-term recurrence ^23,24^. Our model allows discrimination of patients with very high risk within the next 3 months after the index stroke, who may benefit from intensive follow-up, or even more aggressive therapeutic strategies. On the other hand, our model also identifies those patients with a moderate risk of recurrence along the first year after stroke, even with lower accuracy. The discrimination in 3 categories difficults the model prediction, but we considered it relevant to differentiate those patients that had a high recurrence risk within the first year, given that treatment interventions are easier to adopt during this first year after stroke. Furthermore, the individual prediction can be continuously updated according to the patient’s control of modifiable risk factors included in the model. The model is based on easily available variables allowing its use in regular clinical practice. The complete ML model requires more than 40 variables, but the performance of the simplified model remains in the same range.

The most powerful predictive factors for stroke recurrence found in both Cox and ML models were slightly different from those frequently described in the literature. Consultation in an emergency department in the previous 3 months emerged as one of the most powerful predictors of stroke recurrence. It is well-known that a recent previous stroke is a strong predictor of recurrence, and in our ML model, time from previous stroke was the most relevant factor. Also, a non-stroke related consultation in the emergency department was associated with an increased risk of recurrence. Inflammation and infection have been epidemiologically related with IS through several mechanisms ^25,26^, and are among the most common medical consultations in emergency departments in the aged population ^27^.

Anaemia has recently emerged as a relevant cardiovascular risk factor, showing a multifactorial pathophysiological relationship with stroke ^28^. In the Acute Stroke Registry and Analysis of Lausanne (ASTRAL) registry, haemoglobin levels were inversely related with stroke recurrence and mortality ^29^. Atrial fibrillation is a worldwide known risk factor for stroke occurrence ^30^, and even in patients with adequate treatment, the risk of IS recurrence is high ^31^. Furthermore, in these patients, anticoagulation also leads to an increased risk of ICH ^32^.

On the other hand, age was not one of the strongest predictors of stroke recurrence. We detected a high mortality in aged stroke patients which may have diluted the effect on recurrence. In younger patients, increasing age was significantly associated with a higher risk of recurrent stroke.

Another unexpected factor related with stroke recurrence was the Barthel Index (BI), a common scale used to evaluate daily basic activities ^33^. Early BI is a strong prognostic tool for health-care needs, destination at discharge (i.e nursing-home) and functional recovery in stroke patients ^34^. However, no specific relation with stroke recurrence had been previously described. Our hypothesis is that patients with lower BI probably have a worse general health status and sedentary lifestyle, which has been previously associated with an increased risk of stroke, and is one of the targets to address in stroke prevention ^35,36^. Modified Rankin scale (mRS) ^37^ is the most generalised scale to estimate functional status after stroke, but was not available in our public global health database. We plan to test our model in hospital discharge series and will evaluate the mRS as a potential predictor of stroke recurrence.

Regarding modifiable risk factors, all are included in the model, with different weights and relevance. Although the stronger predictors of recurrence in our model are non-modifiable, an adequate modifiable risk factors control showed to significantly decrease the risk of recurrence (up to 62% reduction), as shown in Figure 4D based on our Cox model. Previous studies have shown that patient involvement in their self-care increases the probability of favourable outcomes^38^. PRERISK provides to stroke survivors an accurate prediction tool of their individualised risk of stroke recurrence, and can show graphically how this risk evolves if modifiable risk factors remain under control, which can be used as a motivational strategy to increase the engagement and compliance in secondary prevention treatments ^39^. We have recently developed a web-based platform and mobile app for patient communication and follow-up (NORA, Norahealth, Barcelona, Spain, www.nora.bio)^40^. We plan to integrate PRERISK in the patient’s app, providing updated information of the individual risk of stroke recurrence, aiming to improve stroke awareness and risk factors control.

Prediction of stroke recurrence is a difficult task. To date, the accuracy of the most widely used predictive scores, such as the ABCD2 with diffusion weighted imaging (DWI) ^15^, the Essen Stroke Risk Score (ESRS) ^41^ and Stroke Prognosis Instrument-II (SPI-II) ^42^ range between 0.40 to 0.80 ^13^. Overall, these scores lose accuracy when they are externally validated in different series^13^. Some of these scales, such as the ABCD2, were initially developed with few and easy to acquire variables; however, given the low accuracy, more complex factors were progressively added to increase the predictive power. These additional factors improved the model performance, making the score conversely less generalizable in clinical practice. Furthermore, such scores are mainly restricted to a specific stroke subtype or condition (i.e. TIA or minor strokes) or to a specific range of time after stroke (early or late recurrence) ^13^.

In our study, both ML and Cox models provided fairly good predictions of recurrence risk classes. However, the ML model performs significantly better than Cox model, especially for the early and long-term prediction. One of the advantages of the Cox model is a lower algorithmic complexity. In addition, the outcomes of hazard models are hazard ratios, which represent valuable information for patient communication and risk prevention, as shown in our modelling curves of the individual risk change of a patient according to his vascular risk factors control. However, although the Cox model does not assume any particular “survival model”, it does assume that the effects of the predicting variables upon survival are constant over time and are additive in one scale. Conversely, decision tree models like ADA or RF classifiers analysed here, do not require any assumption or previous patient information and they can be used for categorical classification where cox model becomes infeasible (i.e., prediction of treatment adherence, physical or cognitive complications).

A criticism that emerges with the application of complex ML models is the “black box” effect ^43^: even if the output of the ML model is very accurate, it may not be easily understandable, for instance in terms of the collection of features that have contributed to develop the algorithm ^44^. Several ML predictive strategies ^45^ like the one used in our study or Shapley Additive explanations ^46^ aim to fill this gap, providing good predictions while offering explainable and interpretable models ^47^. Therefore, the PRERISK ML model can be used not only as a predictive tool, but also to identify new relevant factors and novel treatment targets.

The study has several limitations. It is based on public healthcare registries that by their nature have potential bias in data assignment and recollection ^48^. However, the distribution of risk factors, stroke subtypes and rate of recurrence is in concordance with the literature, which provides consistency to our database.

Unfortunately, we do not have data or relevant variables like family medical history of stroke or compliance to treatments. We are aware that preventive stroke treatment compliance plays a critical role for stroke recurrence ^49^. We included data of different drugs prescribed to patients; however, their influence in the models is not clear as there is the lack of information about treatment adherence. Given its plausible relevance, we plan to include treatment adherence in future models. Finally, our data is based on the population of a region of Spain, and reproducibility in other populations needs to be tested.

In conclusion, our predictive model based on ML analysis improves risk prediction and offers individualised information of the probability of stroke recurrence to patients, that can be re-calculated according to their risk factors control. ML based approaches provide statistical advantage for early and long recurrence predictions over classical Cox models. This information can be used as feedback for secondary prevention strategies and enhance patient engagement and treatment compliance. Our results pave the way to scalable ML assisted care to optimise prevention strategies in stroke as well as in other chronic conditions.

## Data Availability

The data that support the findings of this study are available from the corresponding author, [MR], upon reasonable request.

## ACKNOWLEDGEMENTS

We thank Pietro Caliandro (Policlinico Universitario A. Gemelli), Carolina Migliorelli Falcone (Eurecat - Centre Tecnològic de Catalunya), Davide Cirillo (BSC - Barcelona Supercomputing Center) for the fruitful discussions.

## SUPLEMENTAL MATERIAL

- Tables I, II, III and IV.

